# The role of hippocampal theta oscillations in working memory impairment in multiple sclerosis

**DOI:** 10.1101/2020.06.15.20127191

**Authors:** Lars Costers, Jeroen Van Schependom, Jorne Laton, Johan Baijot, Martin Sjøgård, Vincent Wens, Xavier De Tiège, Serge Goldman, Miguel D’Haeseleer, Marie Beatrice D’hooghe, Mark Woolrich, Guy Nagels

**Author notes:** Correspondence, Pleinlaan 2, 1050 Brussels (Belgium).

## Abstract

Working memory (WM) problems are frequent in persons with multiple sclerosis (MS). Even though hippocampal damage has been repeatedly shown to play an important role, the underlying neurophysiological mechanisms remain unclear. This study aimed to investigate the neurophysiological underpinnings of WM impairment in MS using magnetoencephalography (MEG) data from a verbal n-back task. We analysed MEG recordings of 79 MS patients and 38 healthy subjects through event-related fields (ERFs) and theta (4-8 Hz) and alpha (8-13 Hz) oscillatory processes. Data was source reconstructed and parcellated based on previous findings in the healthy subject sample. MS patients showed a smaller maximum theta power increase in the right hippocampus between 0 and 400 ms than healthy subjects (*p* = 0.014). This theta power increase value correlated strongly and negatively with reaction time on the task in MS (*r* = −0.32, *p* = 0.029). Evidence was provided that this relationship could not be explained by a confounding relationship with MS-related neuronal damage. This study provides the first neurophysiological evidence of the influence of hippocampal dysfunction on WM performance in MS.

## Introduction

Cognitive impairment is a significant burden for people with multiple sclerosis (PwMS), having an impact on their employment status, mental health and social and vocational activities. About 43 to 70% of the 2.3 million multiple sclerosis (MS) patients worldwide (Multiple Sclerosis International Federation, 2013) are estimated to suffer from cognitive impairment (Peyser, Rao, LaRocca, & Kaplan, 1990; Rao, Leo, Bernardin, & Unverzagt, 1991) together with the strong physical problems that are caused by the inflammatory demyelination and neurodegeneration that characterise this disorder. Particularly information processing speed (Deluca, Chelune, Tulsky, Lengenfelder, & Chiaravalloti, 2004; Van Schependom et al., 2015) and working memory (WM) (Chiaravalloti & DeLuca, 2008; D’Esposito et al., 1996) but also attention and visuospatial abilities have been shown to be affected in PwMS, even in persons with only mild physical problems (Ruchkin et al., 1994).

Multiple functional magnetic resonance imaging (fMRI) studies have investigated working memory in MS using the n-back task, a task in which a subject has to respond when an item in a sequence is the same as the *n*-th item before. By increasing *n*, the WM load or the number of items to be retained in WM is increased. Those studies have reported relatively heterogenous findings of possibly compensatory mechanisms in different regions (Forn et al., 2007; Penner, Rausch, Kappos, Opwis, & Radü, 2003; Sweet, Rao, Primeau, Durgerian, & Cohen, 2006; Sweet, Rao, Primeau, Mayer, & Cohen, 2004; Wishart et al., 2004). The most common finding for these studies was increased activation in the prefrontal cortex in MS patients (Forn et al., 2007; Sweet et al., 2004). One study also found a decreased activation in the right hippocampus (Sweet et al., 2004). A decreased activation of the hippocampi was also found in an fMRI study of episodic memory in cognitively impaired compared to cognitively preserved MS patients (Hulst et al., 2015). While these fMRI studies were able to point to some specific brain regions possibly involved in WM impairment in MS, they have the disadvantage of low temporal resolution which makes it difficult to draw conclusions about which specific WM processes or mechanisms are disturbed in MS.

Electroencephalography (EEG) n-back studies in healthy subjects reported P300 event-related potentials (ERPs) along the central and parietal electrodes, with amplitudes decreasing with increasing levels of memory load (Ahonen, Huotilainen, & Brattico, 2016; Causse, Peysakhovich, & Fabre, 2016; Dong, Reder, Yao, Liu, & Chen, 2015; Scharinger, Soutschek, Schubert, & Gerjets, 2017) (for review see (Kok, 2001)). Covey and colleagues (Covey, Shucard, & Shucard, 2017) have found that the amplitude of this P300, but also an earlier P100 component, were lower in PwMS than healthy subjects. Such P300 amplitude difference was also already described in an early study, albeit below statistical significance (Ruchkin et al., 1994). Another study found the latency of the N200 and P300 component to be larger in PwMS than healthy subjects, with n-back specific cognitive training causing these differences to disappear (Covey, Shucard, Benedict, Weinstock-Guttman, & Shucard, 2018). A recent magnetoencephalography (MEG) study was able to identify the right inferior temporal and parahippocampal gyrus and the left inferior temporal gyrus as the sources of the n-back M300, the magnetic counterpart of the P300 (Costers et al., 2020).

Rather than ERPs, which are not assumed to be specific to WM, specific frequency band oscillations and their interactions have been shown to be essential for WM function. Theta band oscillations have been thought to be essential for WM activity, organising different WM sub-processes through theta synchronisation (Roux & Uhlhaas, 2014) and encoding the temporal information of WM items in theta oscillatory cycles (Hsieh, Ekstrom, & Ranganath, 2011; Lisman & Idiart, 1995; Lisman & Jensen, 2013; Roberts, Hsieh, & Ranganath, 2013). In a recent study, frontal theta oscillations were causally shown to be excitatory of nature and responsible for prioritising relevant information during WM (Riddle, Scimeca, Cellier, Dhanani, & D’Esposito, 2020). This study also provided causal evidence for the inhibitory influence of parietal alpha band oscillations to suppress WM representations. The function of alpha oscillations during WM has already previously been shown to be modulatory, through mechanisms of inhibition and release from inhibition. This was based on observations of increased neural firing correlating with decreases in alpha power in monkeys (Haegens, Nácher, Luna, Romo, & Jensen, 2011).

During the n-back task, multiple EEG studies in healthy subjects have extensively described increases in frontal theta band power (Dong et al., 2015; Jensen & Tesche, 2002; Kawasaki, Kitajo, & Yamaguchi, 2010) and decreases in occipital-parietal alpha or beta band power compared to baseline (Chen & Huang, 2016; Dong et al., 2015) (for review see (Roux & Uhlhaas, 2014)). A recent MEG study also suggested the orbitofrontal cortex to be the source of this increase in frontal theta power (Costers et al., 2020). Another source of theta power increase was found to be the right hippocampus. While the measurability of the hippocampus using MEG has been heavily disputed, recent evidence has led to more acceptance in the field (see (Pu, Cheyne, Cornwell, & Johnson, 2018) for review). The strongest alpha and beta band power decreases were observed at later timepoints in the right hippocampus and occipital fusiform gyri, involved in letter recognition (Devlin, Jamison, Gonnerman, & Matthews, 2006; James, James, Jobard, Wong, & Gauthier, 2005; Joseph, Gathers, & Piper, 2003; Pernet, Celsis, & Démonet, 2005), suggesting that these regions together enable successful letter encoding and storage in WM. Although the importance of the hippocampus during WM was already established by intracranial observations in rodents and humans (N. Axmacher et al., 2010; Nikolai Axmacher et al., 2007) (for review see (Leszczynski, 2011)) and hippocampal lesioning studies (Fortin, Agster, & Eichenbaum, 2002; Kesner, Gilbert, & Barua, 2002), the findings from the MEG study mentioned above also add to the evidence (Costers et al., 2020). In MS, multiple studies have reported relationships between WM performance and hippocampal atrophy or microstructural damage (R. H.B. Benedict, Ramasamy, Munschauer, Weinstock-Guttman, & Zivadinov, 2009; Koenig et al., 2019, 2014; Longoni et al., 2015; Planche et al., 2017; Preziosa et al., 2016; Sacco et al., 2015; Sicotte et al., 2008). However, no neurophysiological correlates have been reported yet.

We investigated the neurophysiological underpinnings of WM impairment in MS by analysing MEG n-back data of 79 MS patients and 38 healthy controls (HCs). We performed an ERF and time-frequency analysis using the regions-of-interest (ROI) reported using the healthy control data (Costers et al., 2020). In addition to a more traditional group timeseries analysis based on comparing group means, we employed an approach considering individual maximum ERFs amplitudes or power changes within defined time windows. This was motivated by observations of a larger inter-individual latency variation in the MS group when compared with HCs (e.g. see (Covey et al., 2017)). Performing an analysis based on group means could be biased when there is a large difference in heterogeneity between both groups. For example, the mean amplitude of an ERP in a sample with large latency variation would be smaller than in a group with the same individual data but a smaller latency variation (e.g. see (Ouyang, Sommer, & Zhou, 2016)).

Based on previous EEG research (Covey et al., 2017), we hypothesised to observe a lower M300 amplitude in the right and left inferior temporal gyrus. Looking at the results reported by fMRI studies in MS (Forn et al., 2007; Penner et al., 2003; Sweet et al., 2004) that coincide with the regions reported in the n-back healthy control study using MEG (Costers et al., 2020), we expected group differences in the prefrontal regions (Forn et al., 2007; Penner et al., 2003; Sweet et al., 2004), the right fusiform gyrus and right hippocampus (Sweet et al., 2004). Considering the excitatory nature of theta oscillations and the inhibitory nature of alpha oscillations during n-back working memory, as recently shown by (Riddle et al., 2020), and their observation in those regions in (Costers et al., 2020) this would translate in a larger increase of theta power in the prefrontal cortex and a larger decrease in alpha power in the right fusiform gyrus in MS patients. As the hippocampus in the previous HC study (Costers et al., 2020) showed an early increase in theta power followed by a decrease in alpha power, we hypothesised to observe a smaller increase in theta power and/or a larger decrease of alpha power in MS patients compared to HCs.

## Methods and materials

### Participants

We analysed data from 79 MS patients and 38 HCs aged between 18 and 65 years performing a visual verbal n-back task in a MEG scanner. All MS patients were recruited at the National MS Center Melsbroek using the following inclusion criteria: MS diagnosis according to the revised McDonald criteria (Polman et al., 2011) and an Expanded Disability Status Scale (EDSS) (Kurtzke, 1983) score smaller or equal to six at the time of recruitment. Exclusion criteria were having had a relapse or corticosteroid treatment in the six weeks preceding the study as well as having a pacemaker, dental wires, psychiatric disorders or epilepsy. All subjects had normal or corrected vision. The MS population had a mean age of 47.9 ± 9.6 years, education level of 13.9 ± 2.7 years and consisted of 72.2% females (see Table 1). The mean disease duration of the MS sample was 15.1 ± 7.9 years and consisted of 87.3 % or 69 relapsing-remitting MS (RRMS), 8.9% or seven primary-progressive MS (PPMS), 2.5% or two secondary-progressive MS (SPMS) and 1.3% or one clinically isolated syndrome (CIS) patients. The MS patients had a median EDSS score of 3 (inter-quartile range (IQR): [2 – 4]). Of the 79 MS patients, 19 (24%) were treated with benzodiazepines. The subjects in the HC sample had a mean age of 48.0 (± 12.0) years, an education level of 15.0 ± 2.0 years and was comprised of 60.5 % females. Years of education was counted from the start of primary school in Belgium (e.g. 12 years = finished middle school). All participants provided written informed consent. Ethical approval for the study was provided by the ethics committees of the National MS Center Melsbroek and the University Hospital Brussels (Commissie Medische Ethiek UZ Brussel, B.U.N. 143201423263, 2015/11).

**Table 1.**
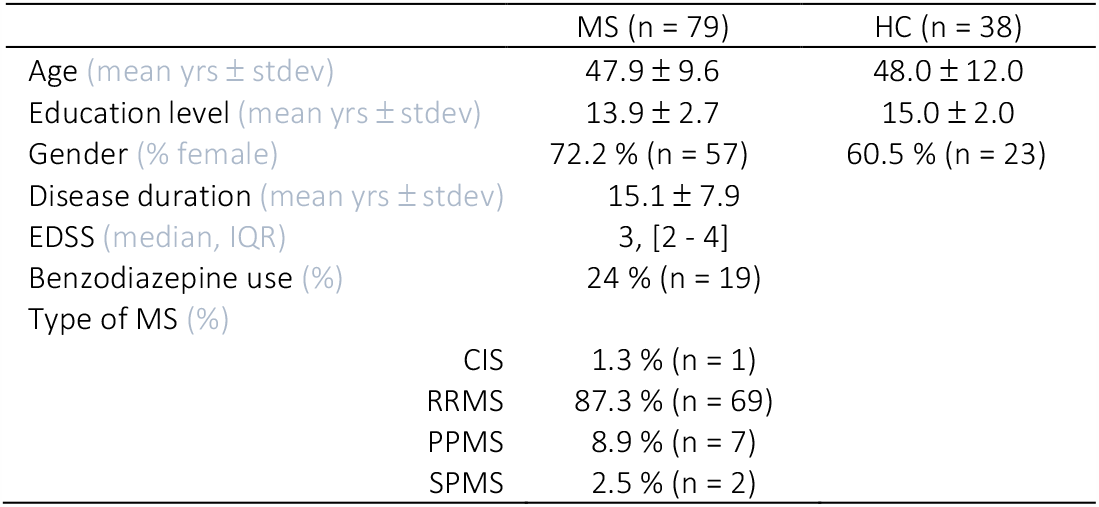
Demographic and clinical characteristics of the MS and HC sample. EDSS = Expanded Disability Status Scale; HC = healthy controls; MS: multiple sclerosis; CIS = clinically isolated syndrome; RRMS = relapsing-remitting MS; PPMS = primary progressive MS; SPMS = secondary progressive MS; yrs = years.

### Paradigm

We employed a visual verbal version of the n-back paradigm with three conditions or levels of WM load (0, 1 and 2-back). The task was split into 4 blocks per condition, with a total of 12 blocks which were presented in a pseudo-random order. Every block consisted of 20 stimuli, for a total of 240 stimuli and respectively 25, 23 and 28 target trials per condition. Subjects were asked to respond with a button press using their right hand when the letter that appeared on the screen was the letter X (0-back), the same letter as the one before in the sequence (1-back) or the same letter as two letters before (2-back). The paradigm is illustrated in Fig. 1. The letter stimuli had a size of 6 by 6.5 cm and were projected on a screen 72 cm from the front of the MEG helmet. Each stimulus was presented on the screen for 1 second, with a 2.8 second inter-trial interval. The instructions for the specific conditions were presented at the start of every block for 15 seconds. The onset of the visual stimuli was measured using a photodiode. Importantly, we also included non-target trials in our analysis as the cognitive processes we are interested in are the stimulus processing, WM encoding and storage which is performed in every single trial. An additional reason was the resultant substantial increase of the number of trials included in the analyses. Trials during which an incorrect button press was recorded were removed from the analyses. Per subject, we removed spurious outlier reaction times using a conservative cut-off of three or more scaled median absolute deviations. Importantly, we only analysed MEG data from the 2-back condition because the involvement of WM in the 0 and 1-back condition are highly disputable.

**Figure 1.**
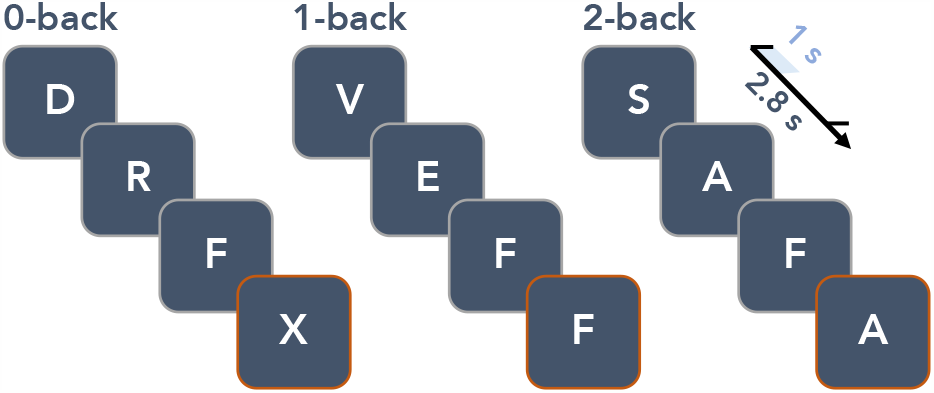
Illustration of the visual verbal n-back paradigm. The inter-trial interval was 2.8 s and stimuli were presented for 1 s on the screen. Target stimuli which required a response are highlighted with an orange border.

### Data acquisition

The MEG acquisition for the first 40 subjects was performed using an Neuromag VectorView™ system, while the last 77 subjects were scanned using an Neuromag™ TRIUX system (MEGIN Oy, Croton Healthcare, Helsinki, Finland) due to a system update at the CUB Hôpital Erasme (Brussels, Belgium). Both whole-head systems use 306 channels, of which 204 planar gradiometers and 102 magnetometers, and are placed in a lightweight magnetically shielded room (MSR, Maxshield™, MEGIN Oy, Croton Healthcare, Helsinki, Finland). The characteristics of the MSR have been described elsewhere (De Tiège et al., 2008). Participants were instructed to sit as still al possible during the acquisition and were seated in an upright position with their head positioned to the back of the MEG helmet. An electrocardiogram (ECG) and horizontal and vertical electrooculogram (EOG) was recorded to be used in offline artefact rejection. Four head position indicator coils were also attached to the left and right forehead and mastoid to track head movement. In order to allow qualitative co-registration with the subjects’ 3D T1-weighted anatomical magnetic resonance image (MRI) we registered a minimum of 400 points on the scalp and nose using Polhemus FASTTRAK 3D digitizer (Polhemus, Colchester, VT, USA) together with three fiducials (nasion, left and right preauricular), to obtain the subjects’ head shape. The MR image was collected using a 3 Tesla Philips Achieva scanner (Amsterdam, Netherlands) located at the Universitair Ziekenhus Brussel (Jette, Belgium). The data for this study are not publicly available. Researchers interested in a collaboration on these data are welcome to contact the senior authors. Analysis scripts are available upon request from the corresponding author.

### Data pre-processing and source projection

The first step of data pre-processing was using the temporal extension of the signal-space separation algorithm implemented in the Maxfilter™ software (version 2.2 with default parameters; MEGIN Oy, Croton Healthcare, Helsinki, Finland) to reduce external noise and correct for head movements. All the next steps of pre-processing were performed using Oxford’s Software Library (OSL; Oxford Centre for Human Brain Activity, U.K., https://ohba-analysis.github.io/osl-docs/), a software library designed for analysing MEG data using functions from SPM12 (Welcome Trust Centre for Neuroimaging, University College London) and Fieldtrip (Oostenveld, Fries, Maris, & Schoffelen, 2011). Using OSL, the data were converted to SPM format and downsampled to 250 Hz. Independent component analysis with automatic artefactual component rejection was then performed using OSL’s AFRICA function. Rejections of components were based on correlations with EOG and ECG recordings (> 0.5) and kurtosis (> 20). The resulting signal and rejected components were manually checked for every participant. Next, data were high-pass filtered at 0.1 Hz, co-registered with individual MRIs, and epoched (see below). Using an auto-kick detection in the eigenspectra we identified the number of principal components in the data which was reduced after Maxfiltering. Within those PCs the minimum eigenvalue was used to normalise the different sensor types. We rejected bad channels and trials using generalised extreme studentized deviate tests (GESDs) (Rosner, 1983), with a cut-off of significance at 0.05. We excluded an average of 5.9 ± 6.7 and 4.5 ± 5.7 % of channels in the, respectively, MS and HC sample. An average of 5.2 ± 4.6 (MS) and 5.9 ± 4.7 % (HC) of trials were abstained from analysis. Finally, an average of 3.8 ± 2.2 (MS) and 3.2 ± 1.2 (HC) components were rejected. Source reconstruction was performed using a bilateral beamformer employing Bayesian principal component analysis (PCA) (Woolrich, Hunt, Groves, & Barnes, 2011). This approach uses a data-driven estimate of the data covariance matrix that automatically trades-off between the signal-to-noise and spatial resolution (Woolrich et al., 2011). A single-shell forward model was used, with a projection on a 5 mm dipole grid.

### Parcellation

We parcellated the data using a downsampled version of the Harvard-Oxford (sub-)cortical Atlas. The probability-based atlas with 1 mm^3^ resolution was thresholded at 25% probability and then downsampled using FSL’s FLIRT function to 5 mm^3^ resolution. After this, the downsampled atlas was again thresholded at 25% probability to produce an unweighted parcellation atlas. In OSL, the parcel timeseries were calculated by extracting the 1^st^ principal component from all voxel timeseries in the parcel. We selected the regions of interest – frequency band pairs based on the findings in the healthy control analyses (Costers et al., 2020) (see Fig. 2 for schematic representations of the parcellation atlases). Important to note is that we included ROIs found in healthy controls to be a local maximum in any of the three n-back conditions, while we only analyse data from the 2-back condition in this study. This was done in order to avoid not including relevant ROIs which could have been narrowly missed as a local maximum in the 2-back condition. For the theta band we included the precuneus, right frontal orbital, left frontal orbital, right middle frontal, right thalamus, right hippocampus and right inferior temporal. For the alpha band, we included the right thalamus, right hippocampus, right temporal occipital fusiform, left occipital fusiform, left hippocampus and left superior parietal. As we assumed focality of the sources of ERFs, based on observations in the HCs (Costers et al., 2020), we used spherical ROIs with a diameter of 12mm around the voxel of maximum effect on group level for our ERF analysis. We used anatomical parcellations for the analysis of time-frequency effects because these were less focal, making the interpretation and relation to previous findings easier. We included only the ROIs reported in (Costers et al., 2020) assumed to be relevant for cognition, being the precuneus and the right and left inferior temporal gyrus, thus excluding the cerebellum and sensorimotor network from these analyses. The coordinates for these three ROIs were [10 −75 52], [40 −20 −32] and [-40 0 47] mm in MNI space.

**Figure 2.**
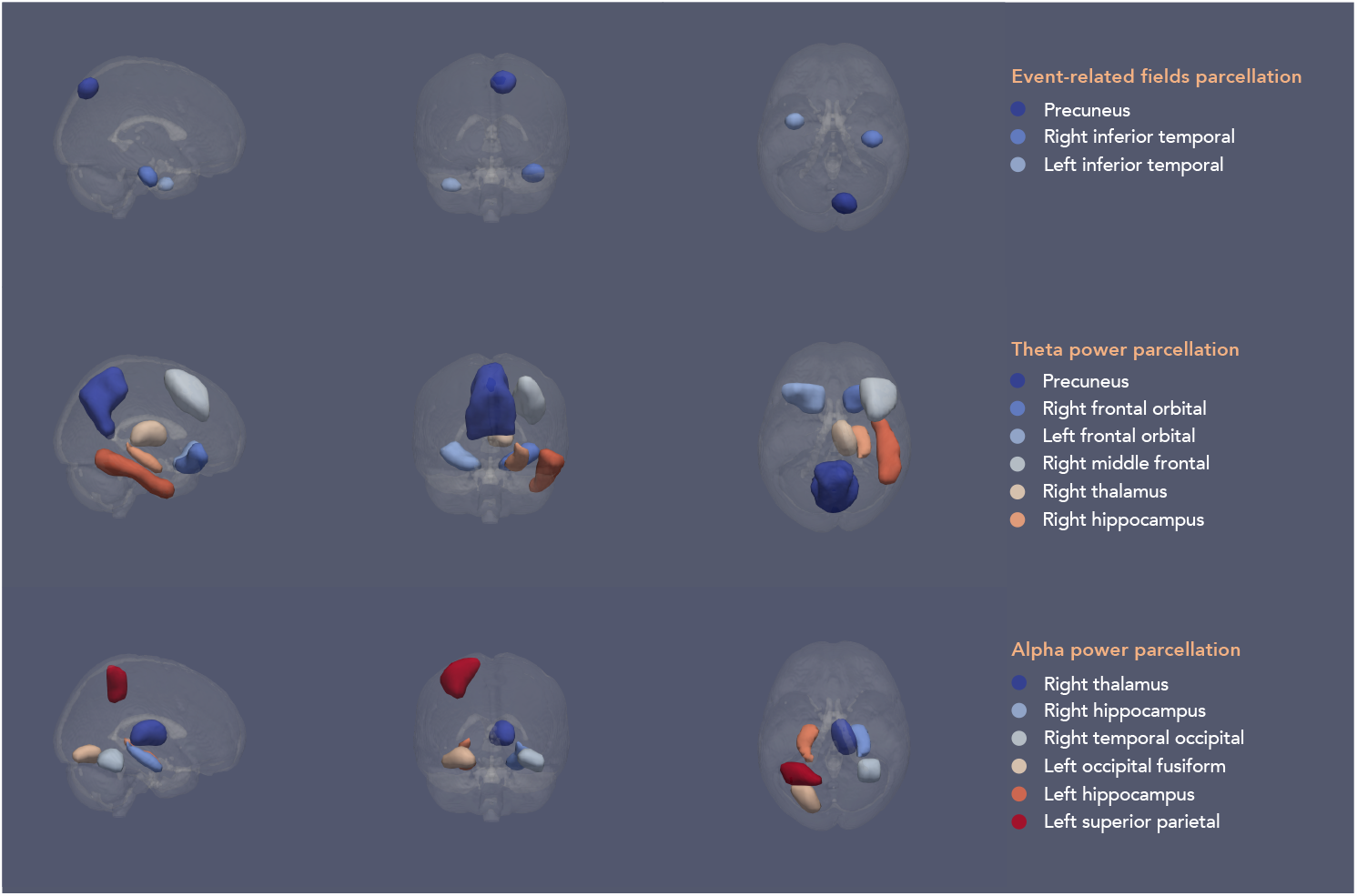
Schematic of the parcellation atlases for the different analyses based on findings in healthy controls (Costers et al., 2020). The parcels in the schematic have been smoothed solely for illustrative purposes. Parcellation atlases using 5mm^3^ voxels were used for the analyses

### Event-related field analysis

OSL was used to perform source reconstruction, parcellation and first-level analysis. For the ERF analysis, data was filtered between 0.1 and 40 Hz and epoched from [-200 – 800] ms relative to stimulus onset. Baseline correction using the time window −200 ms until stimulus presentation was performed in Fieldtrip, after which subject-level and group-level analysis was performed. As mentioned before, we performed two types of analyses. A more traditional group EFR analysis using MaxStat correction (Nichols & Holmes, 2002) over all parcels and timepoints (see Statistics), and an analysis based on individual maximum ERF amplitudes. For our maximum peak value group analysis we based the time windows of interest on the healthy control sample, resulting in a time window of [50 – 250] ms for the precuneus, [300 – 700] ms for the right inferior temporal and [150 – 400] ms for the left inferior temporal gyrus.

### Time-frequency analysis

For time-frequency analysis, the data was filtered between 0.1 and 80 Hz. In order to avoid edge artefacts, epochs were initially kept at −1.25 s to +1.55 s relative to stimulus onset. The fixed inter-trial interval was 2.8 s, so there was no overlap of epochs. Time-frequency analysis was performed using the Morlet wavelet method (6 cycles) as implemented in Fieldtrip. We used a 0.5 Hz frequency resolution in the theta [4 – 8] Hz and alpha [8 – 13] Hz band over which the data was then averaged. Baseline normalisation was performed using decibel conversion (dB = 10*log_10_(power/baseline)) with a −500 to −100 ms baseline period, after which data was epoched to −200 to 800 ms to remove edge artefacts. Subject-level and group-level analysis were performed in Fieldtrip. As mentioned before, we performed a group analysis using MaxStat correction (Nichols & Holmes, 2002) over all timepoints and parcels (see Statistics). Importantly, this correction was performed per band. The minimum or maximum peak value of theta power increase and alpha power decrease relative to baseline was calculated using the, respectively, time windows [0 – 400] ms and [200 – 600] ms.

### Correlations with performance, clinical and volumetric data

We calculated Pearson correlations between the maximum peak values that showed significant group differences, namely the maximum theta power value in the right hippocampus, and performance on the task (accuracy and reaction time) and clinical parameters (disease duration and EDSS) in the MS sample. In order to gain more insight into our findings and avoid possible confounders we also correlated the maximum theta power increase values in the right hippocampus to the normalised volume of the right hippocampus and scores on the California Verbal Learning Test-II (CVLT-II). The former were calculated from MR images of the MS patients using the icobrain (Jain et al., 2015) (Version 3.1; formerly known as MSmetrix). The latter is a verbal working memory task which has shown high sensitivity for MS (Strober et al., 2009). In addition, we performed post-hoc correlations between median reaction time and Symbol Digit Modalities Test (SDMT) score, a test for information processing speed in MS (Ralph H B Benedict et al., 2012), and normalised white matter volume in order to look into the effects of possible confounders.

### Statistics

For the analysis of the performance data, we used non-parametric Wilcoxon-rank sum tests using the Benjamini-Hochberg procedure to control the false discovery rate (FDR) (Benjamini & Hochberg, 1995). For MEG data, our statistical analyses consisted of a non-parametric maximum statistic (MaxStat) approach (Nichols & Holmes, 2002) based on group permutations correcting for multiple comparisons over all parcels and timepoints of the ERF timeseries or time-frequency power timeseries. Group permutations were performed using random label swapping and Welch t-tests for unequal variances. For a quality check and comparison with results reported in (Costers et al., 2020), we also performed single-group maximum statistic permutation tests. In this case, a dependent t-test was used in the MaxStat approach and the participants’ data was permuted with zero values. This corresponds to a sign flipping procedure which is more standard but currently not implemented in Fieldtrip. All tests were two-tailed, except the tests on performance data, performed in Fieldtrip with 5000 permutations and alpha = 0.05. For the analyses of the ERF and time-frequency maximum peak values, we also used a similar MaxStat approach but using a non-parametric Wilcoxon Rank Sum test. All reported p-values for correlations were controlled for multiple comparisons using FDR correction (Benjamini & Hochberg, 1995). Importantly, before all statistics we regressed out the effects of scanner type (over the whole sample) and benzodiazepines (within the MS sample) from the data.

## Results

### Behavioural data

For the 0-back condition the median reaction time for the HC sample was 423 ms (IQR: [367 – 505]) and for the MS sample 473 ms (IQR: [435 – 509]). For the 1-back condition, the median reaction times for both samples were, respectively, 423 ms (IQR: [361 – 521]) and 483 ms (IQR: [449 – 538]). For the 2-back the median reaction time for the HC sample was 511 ms (IQR: [403 – 575]) and 560 ms (IQR: [488 – 627]). The median accuracy for the 0-back condition for the two groups were both 100% (IQR: [100 – 100]). For the 1-back they were 95.7% (IQR: [95.7 – 100]) and 100% (IQR: [91.3 – 100]) for, respectively the HC and MS group. For the 2-back the median accuracy for the HCs was 89.7% (IQR: [79.3 – 96.6]) and 82.76% (IQR: [55.2 – 93.1]) in MS. We observed a significant difference between the two groups in reaction time on the 0-back (z = - 2.50, p = 0.007), but not in accuracy (z = 0.79, p = 0.215). In the 1-back condition we found both a significant difference between MS patients and healthy controls in reaction time (z = −2.81, p = .007) and accuracy (z = 2.81, p = 0.007). In the 2-back condition we also found a significant group difference in reaction time (z = −2.61, p = 0.007) and accuracy (z = 2.60, p = 0.007). The performed tests were one-tailed Wilcoxon rank sum tests, all p-values mentioned above were corrected for multiple comparisons using false-discovery rate control (Benjamini & Hochberg, 1995).

### Event-related fields

Results did not show significant ERF differences in the three ROIs between the MS and control sample assessed via a MaxStat permutation approach correcting over parcels and timepoints (see Figure 4, left column). See additional Figure 1 for the values of the t-statistics at all timepoints. A statistical comparison of the maximum peak values for the ERFs in the precuneus (z = 1.14, p = 0.32), right inferior temporal (z = 1.28, p = 0.26) or left inferior temporal gyrus (z = 0.38, p = 0.72) also did not show any significant differences between the two groups (see Figure 4, right column). All p-values were corrected for multiple comparisons (see Statistics).

**Figure 3.**
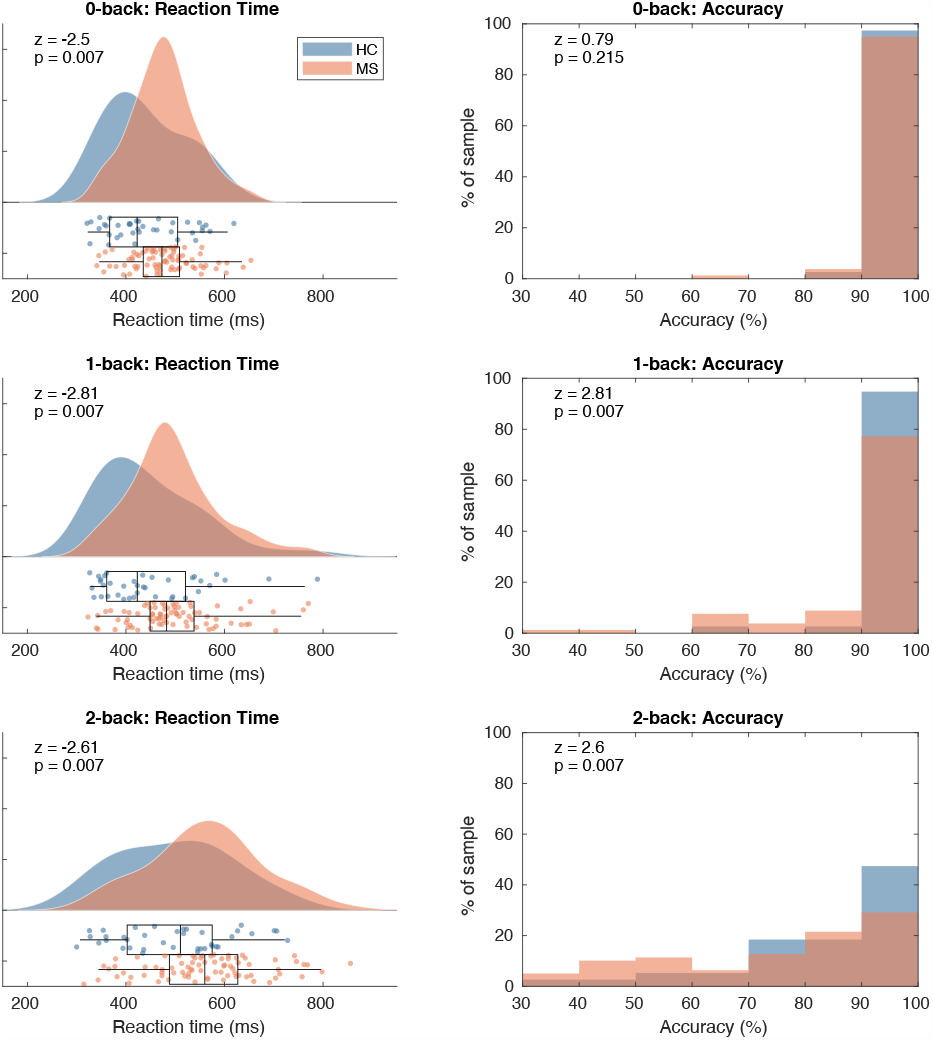
Left panels: reaction time kernel density distribution and boxplots for the three WM load conditions. Right panels: histograms of accuracies per subject. The two groups did not significantly differ in reaction time or accuracy. ERF = Event-Related Field; MS = multiple sclerosis patients; HC = healthy controls.

**Figure 4.**
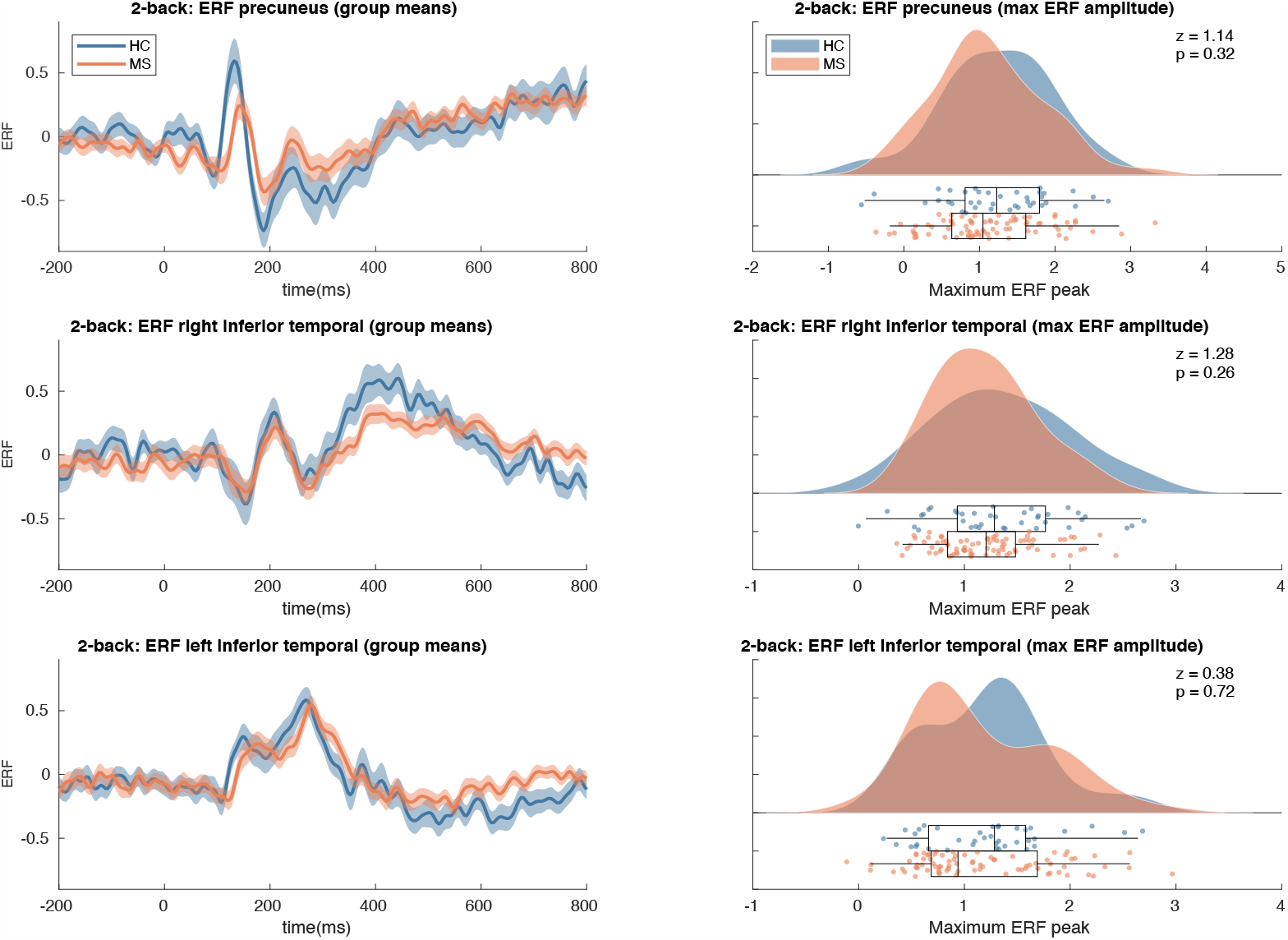
Event-related field results. Left panels: shaded error bars (using standard error) of group mean ERFs in the selected ROIs. Right panels: raincloud plot of distribution of the maximum peaks of ERFs in the selected ROIs. See Additional Figure 1 for t-statistics of the single group and group difference tests on the ERF timeseries. MS = multiple sclerosis patients; HC = healthy controls.

### Time-frequency results

We did not find group differences using the MaxStat approach over the complete timeseries of power changes relative to baseline in any of the twelve theta band or alpha band regions of interest (see Figure 5, left column). The analyses of group differences in the maximum power increase in the theta frequency band showed a significant difference in the right hippocampus (z = 2.98, p = 0.014) but not in the precuneus (z = 2.04, p = 0.21), right frontal orbital (z = 2.15, p = 0.16), left frontal orbital (z = 0.19, p = 0.99) or right thalamus (z = 1.96, p = 0.25) (see Figure 5, right column). In the alpha band no group differences were found in the maximum power decrease in the right thalamus (z = 0.195, p = 0.25), right hippocampus (z = 1.02, p = 0.86), right temporal occipital fusiform (z = 0.37, p = 0.99), left occipital fusiform (- 0.19, p = 0.99), left hippocampus (z = 0.21, p = 0.99) or left superior parietal (z = 1.17, p = 0.78). All p-values were corrected for multiple comparisons (see Statistics).

**Figure 5.**
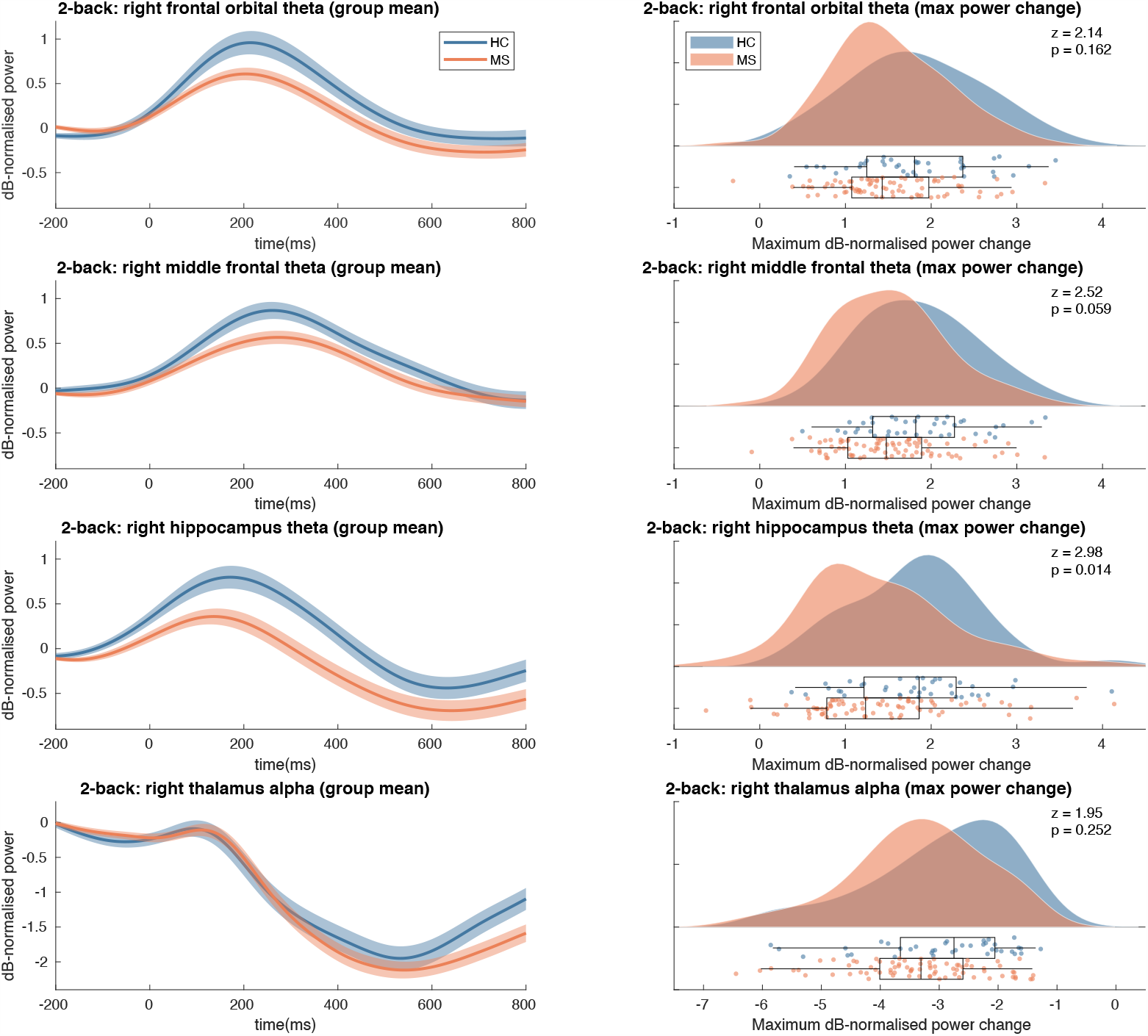
Time-frequency max power changes results. Left panels: shaded error bars (using standard error) of group mean power changes in the selected ROIs. Right: raincloud plot of distribution of maximum power change values per group in selected ROIs. See Additional Figure 1 for t-statistics of the single group and group difference tests on the power change timeseries for all parcels. MS = multiple sclerosis patients; HC = healthy controls.

### Relationship with performance and clinical variables

In order to gain insight in the possible causes of impaired n-back performance in MS, we investigated correlations of the parcel-frequency pair that showed significant group differences (maximum theta band power change in the right hippocampus) and multiple variables such as performance on the task (median reaction time and accuracy), clinical variables (disease duration and EDSS) and score on the CVLT-II verbal WM test (see Fig 6.). We observed a significant correlation between the maximum theta power increase in the right hippocampus and the median reaction time (r = − 0.32, p = 0.029) implying that subjects with a higher maximum theta power increase report a shorter median reaction time. Correlations between the maximum theta power increase in the right hippocampus and 2-back accuracy (r = − 0.21, p = 0.172), EDSS score (r = − 0.2, p = 0.172) and CVLT-II score (r = 0.19, p = 0.172) were not significant. Correlations with disease duration (r = − 0.06, p = 0.634) and normalised right hippocampal volume (r = 0.06, p = 0.634) were small and not significant. Finally, we wanted to gain insight into the possible confounder of general neuronal damage which could explain a delay in reaction time due to damage to white matter tracts and a locally decreased theta response in the right hippocampus. We looked for correlations between the median reaction time and SDMT score (r = − 0.08, p = 0.634), a neuropsychological measure for processing speed in MS, and normalised white matter volume (r = − 0.08, p = 0.634), but both showed to be very weak and not significant. We also explored the correlations mentioned above relevant for HCs but found none to be significant (see Additional Fig. 2). Interestingly, the correlations between median reaction time and SDMT score (r = −0.3, p = 0.203) and normalised volume of white matter (r = −0.37, p = 0.14) were not significant in the HC sample.

**Figure 6.**
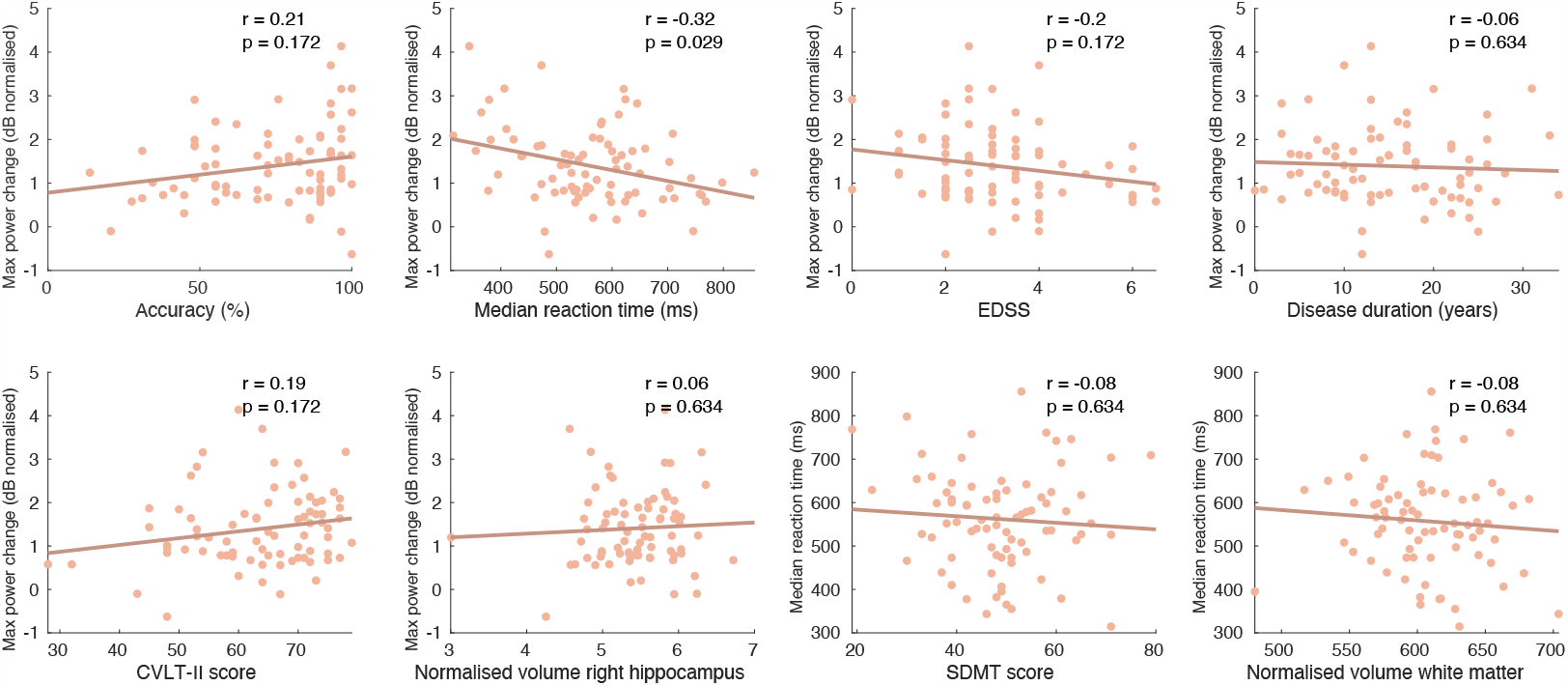
Correlations between the maximum theta power change value in the right hippocampus and performance measures, clinical parameters and volumetric measurements in the MS sample. The last two figures (bottom right) display correlations between the median reaction time and SDMT score and normalised white matter volume. EDSS: Extended Disability Status Scale; CVLT-II: California Verbal Learning Test II; SDMT: Symbol Digit Modalities Test.

## Discussion

This study aimed to investigate the neurophysiological mechanisms of WM impairment in MS. We selected ROIs for an ERF and time-frequency analysis based on results from our HC sample (Costers et al., 2020). A group means based approach was performed as well as an analysis based on individual maximum ERF amplitudes or power changes in the theta and alpha band. Finally, we related parameters that were significantly different between MS patients and healthy controls to performance, and clinical and anatomical volume parameters.

MS patients performed significantly worse in accuracy and reaction time on all conditions, except accuracy in the 0-back condition. Only one of the previous n-back fMRI studies (Forn et al., 2007; Penner et al., 2003; Sweet et al., 2006, 2004) was able to find significant differences in performance between the HC and MS sample (Wishart et al., 2004). A previous EEG study (Covey et al., 2017) and two neuropsychological n-back studies (Covey, Zivadinov, Shucard, & Shucard, 2011; Parmenter, Shucard, Benedict, & Shucard, 2006) did also report prolonged reaction times during the n-back task in MS. We found no group differences in the ERFs in the precuneus, right inferior temporal and left inferior temporal. Unlike a previous scalp EEG study (Covey et al., 2017), we did not find the amplitude of the P300-resembling ERFs to be lower in the MS sample compared to the healthy controls. When comparing the ERF peak amplitudes, which we assume to be a more sensitive approach, we also did not observe a group difference. Possibly, the summative nature of potentials measured on the EEG scalp level leads to ERPs that are more sensitive to small but widespread changes. As an example, a similar effect of a lower P300 amplitude due to an increasing n-back WM load has been repeatedly found at EEG scalp level (Ahonen et al., 2016; Causse et al., 2016; Dong et al., 2015; Scharinger et al., 2017) but has been failed to be reported at source level (Costers et al., 2020).

We did observe a significantly smaller maximum theta band power increase in the right hippocampus in MS patients between 0 and 400 ms. This finding is in line with our hypothesis based on a previous finding of decreased activation in the right hippocampus in MS patients in an n-back fMRI study (Sweet et al., 2004). We based this hypothesis on recent evidence for the excitatory function of theta oscillations during WM (Riddle et al., 2020), assuming that a smaller increase in theta power would translate to the lower activations observed using fMRI. However, such translations of findings from fMRI to MEG are not always as straight-forward and caution must be taken (see (Hall, Robson, Morris, & Brookes, 2014) for review). Hippocampal theta oscillations during WM have been found to play a role in the encoding of temporal information of WM items through observations of cross-frequency coupling using intracranial EEG (iEEG) (Nikolai Axmacher et al., 2010). A recent study using high-density and intracranial EEG as well as fMRI datasets was able to show that in episodic memory the hippocampus functions as the switchboard between perception and memory processes, but that function was related to hippocampal alpha processes taking place after 500 ms (Treder et al., 2020). Due to the parallel nature of WM processes during the n-back task, in contrast to the Sternberg task which allows isolating processes such as WM maintenance, it is difficult to make any claims about the specific processes that reflect this impaired hippocampal theta oscillatory response.

Interestingly our data showed that that this increase in theta power in the right hippocampus had a strong negative correlation with reaction time on the task in MS patients. Thus, MS patients with a less strong hippocampal theta power increase responded slower to the 2-back trials. This suggests that impaired WM function in MS could be related to disturbed theta oscillatory processes in the right hippocampus. A possible confounding common cause for this relationship between the maximum theta power increase and reaction time could be general MS-related neuronal damage. Damage to white matter tracts and a subsequent decrease in conduction velocity, and local (micro)structural damage to the hippocampus are hallmarks of MS neuropathology.

The first two could lead to a longer reaction time while the latter could lead to impaired local theta generation in the hippocampus. In order to test this hypothesis, we calculated post-hoc correlations between the median reaction time and both SDMT score, a measure for information processing speed in MS, and normalised white matter volume, which should reflect white matter damage. Both correlations showed to be very small, suggesting that our finding is not due to the confounding factors of overall widespread white matter atrophy and a general decrease in information processing speed. A correlation between the maximum hippocampal theta power change and hippocampal volume also showed to be small and non-significant. As previously stated, the interpretation of the possible mechanisms that underlie this relationship between impaired hippocampal theta responses and reaction time is challenging, due to the complex functions that the hippocampus has been related to during WM. Future studies should try to investigate the exact WM processes that underly this critical hippocampal theta response.

As mentioned before, it is not surprising to find the hippocampus to play an important role in MS-related WM impairment. It is one of the brain regions most frequently affected by atrophy in MS, which can already be present in clinically isolated syndrome (CIS) (Planche et al., 2018). Hippocampal atrophy or microstructural damage has been repeatedly related to impaired (working) memory function in MS (R. H.B. Benedict et al., 2009; Koenig et al., 2019, 2014; Longoni et al., 2015; Planche et al., 2017; Preziosa et al., 2016; Sacco et al., 2015; Sicotte et al., 2008). For example, a recent study found CA1 atrophy at the time of CIS to be related to episodic verbal memory performance one year after CIS (Planche et al., 2018). Besides the findings of decreased activations during 2-back trials by Sweet and colleagues (Sweet et al., 2004), only two fMRI studies have provided neurophysiological evidence for impaired hippocampal functioning in MS. Roosendaal and colleagues (Roosendaal et al., 2010) observed decreased resting-state connectivity between the hippocampi and the cerebellum, the anterior cingulate gyrus, thalamus, and prefrontal cortex in MS patients. Increased connectivity between the left hippocampus and the right posterior cingulate was reported in MS patients during rest by Hulst and colleagues (Hulst et al., 2015). They also observed lower hippocampal activation during an episodic memory task in cognitively impaired versus cognitively preserved MS patients, which was related to memory status. The findings in this study are novel in that specific neurophysiological evidence of hippocampal WM dysfunction in MS, in this case disturbed theta oscillatory processes between 0 and 400 ms post-stimulus, can be linked to impaired WM performance. Importantly, this finding, and a lack of such finding using an approach based on group means, highlights the importance of correctly characterising a person’s neurophysiological response in a clinically heterogeneous disease as MS, instead of implicitly assuming that the same neural processes are active in all subjects at a certain timepoint.

This finding could potentially lead to the development of new therapies for the improvement of WM in MS. In the context of a recent TMS study that was able to increase WM capacity by applying theta TMS stimulation to the prefrontal cortex or alpha frequency stimulation to the parietal cortex (Riddle et al., 2020), future studies could explore similar stimulation approaches based on the findings from this study. While hippocampal stimulation can be challenging, a recent study showed that after TMS stimulation to a region interconnected to the hippocampus, the posterior inferior parietal cortex, healthy subjects showed improved associative memory formation (Tambini, Nee, & D’Esposito, 2017). The short-lived nature of WM theta processes however, as shown in Fig. 4, should be taken into consideration as well as the succession by alpha oscillatory processes, as observed in the right hippocampus. Considering the inhibitory nature of alpha oscillations during WM processes, there seems to be only a small window of time during which hippocampal processes are necessary for n-back WM activity. This would be a challenge for possible WM therapies in MS based on theta-band TMS stimulation of the right hippocampus.

We did not find any group differences in theta band power changes in the selected ROIs (see Fig. 2) using our MaxStat approach correcting over ROIs and all timepoints. A general trend in the theta band ROIs was that MS patients showed smaller increases in theta power compared to HCs, for example in line with our hypothesis in the right hippocampus, but none of the group differences were significant. In the frontal ROIs this observation seems to be the opposite of our hypothesis of a stronger theta response in MS patients based on previous observations of increased activations in the prefrontal cortex reported by most fMRI studies (Forn et al., 2007; Penner et al., 2003; Sweet et al., 2006, 2004), and causal evidence for the excitatory role of frontal theta oscillations (Riddle et al., 2020). One fMRI study did report less activation in the prefrontal and specifically also in the middle frontal regions in MS patients (Wishart et al., 2004). More neurophysiological M/EEG studies are needed to elucidate the changes in frontal theta oscillations during WM in MS.

In all alpha band ROIs MS patients generally showed a stronger alpha power decrease relative to baseline compared to healthy controls, in line with our hypothesis in the right hippocampus, but none of the ROIs yielded significant group differences at any of the timepoints. In the right fusiform gyrus, this trend was also in line with our hypothesis which was based on causal evidence of the inhibitory role of alpha oscillations during WM (Riddle et al., 2020) and decreased activations in the right fusiform gyrus in MS patients reported during an n-back fMRI study (Sweet et al., 2004). A possible explanation for these non-significant findings could be that the MaxStat procedure which corrects for multiple comparisons over all parcels and timepoints is considered as being relatively conservative. We chose this technique because it allows clear inference about the location of found effects in time and space, in contrast to more sensitive methods such as threshold-free cluster enhancement (Smith & Nichols, 2009).

## Conclusion

This study is the first to provide neurophysiological evidence of impaired hippocampal WM processing in MS that can be related to WM performance. Our data suggests that impaired theta processes in the right hippocampus lead to slower reaction times on the task, which cannot be accounted for by MS-related white matter damage and subsequent decreases in information processing speed.

## Data Availability

The data for this study are not publicly available. Researchers interested in a collaboration on these data are welcome to contact the senior authors. Analysis scripts are available upon request from the corresponding author.

## Acknowledgements

We would like to thank the participants for their time and commitment to this study. We would also like to thank Ann Van Remoortel for her help with the recruitment of participants.

---

**Additional Figure 1.**
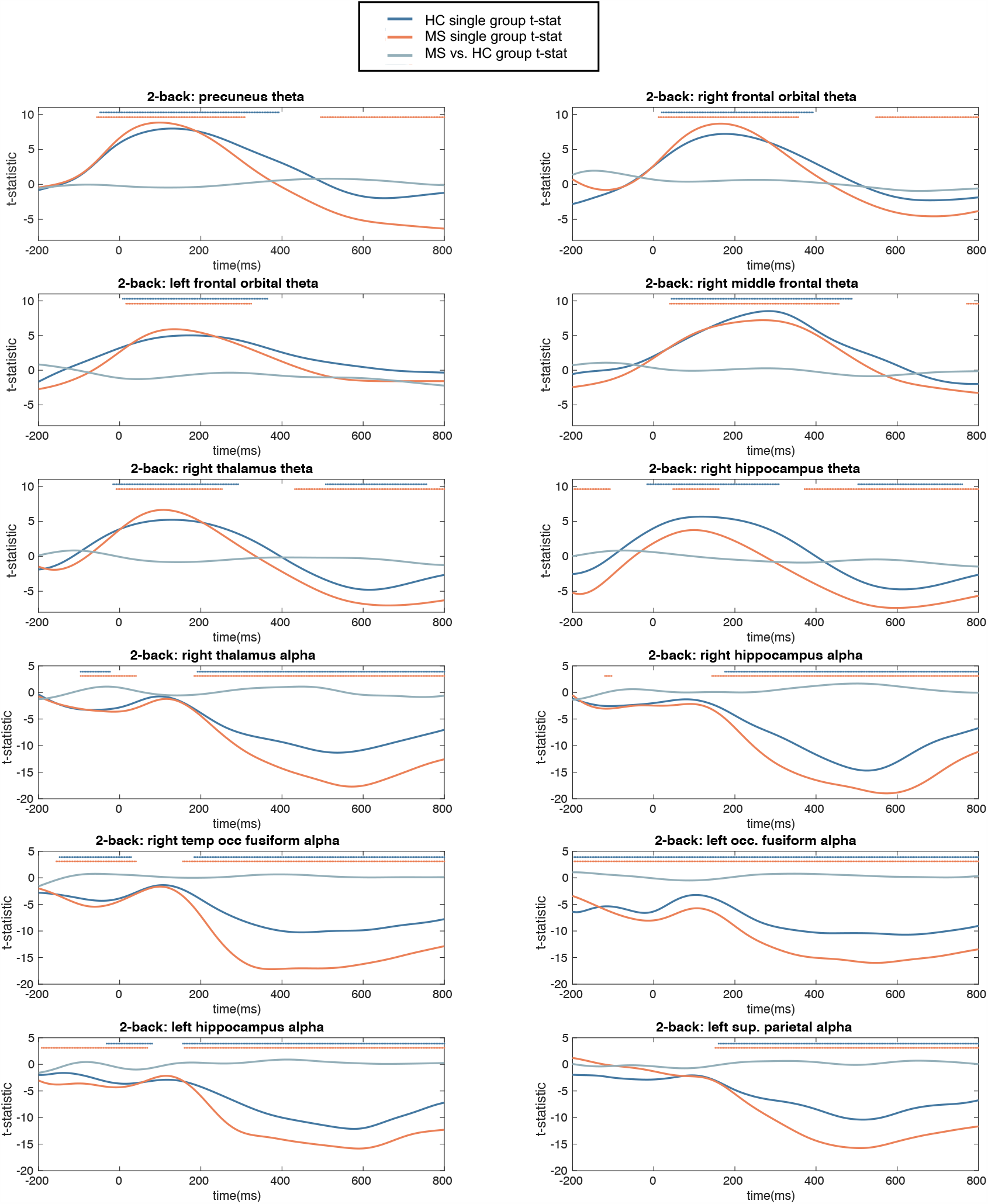
T-statistics of the single-group and group-difference tests on theta and alpha band power. Stars illustrate the significant timepoints calculated using MaxStat correction over parcels (per band) and timepoints.

**Additional Figure 2.**
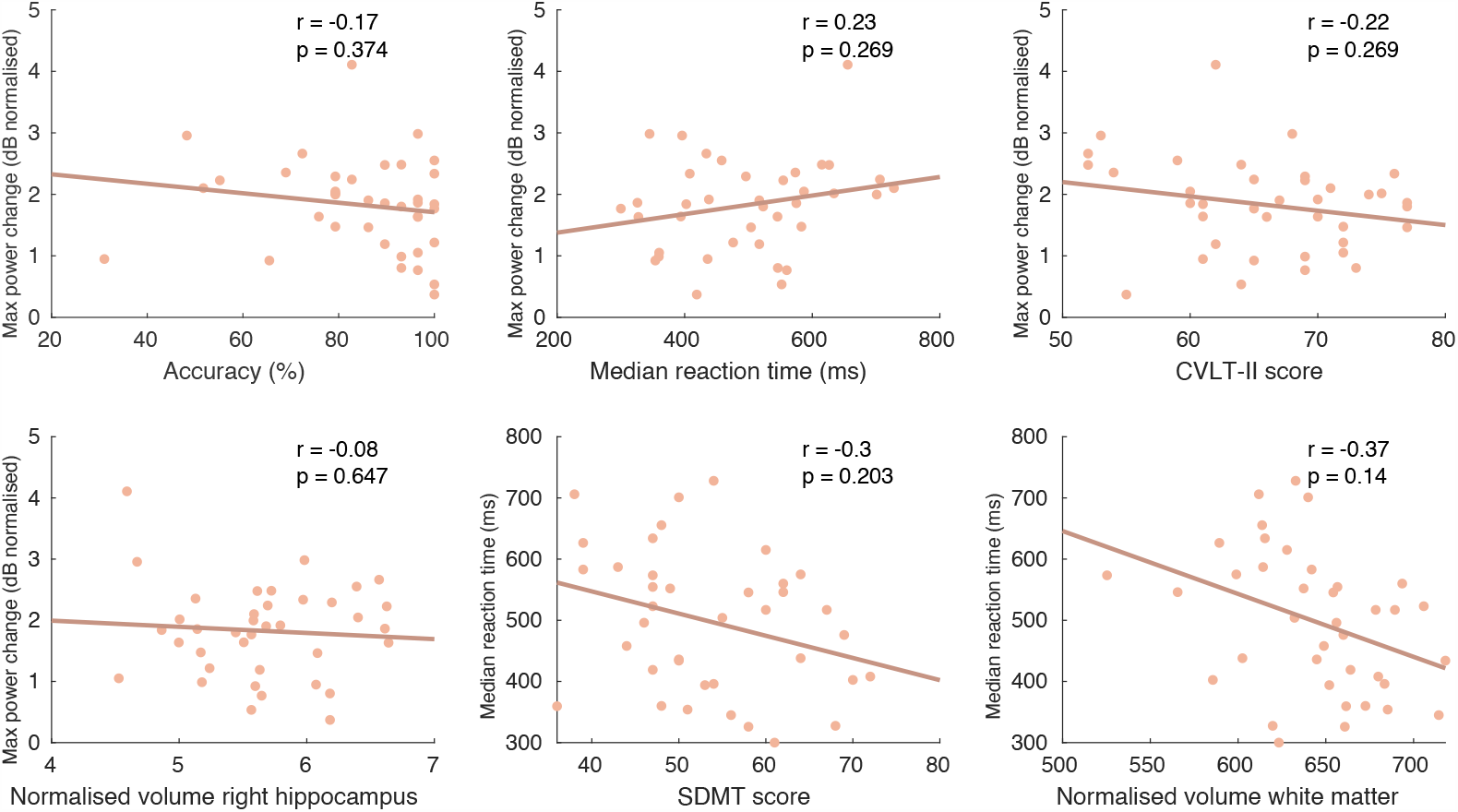
Correlations between the maximum theta power change value in the right hippocampus and performance measures, neuropsychological scores and volumetric measurements in the healthy control sample. CVLT-II: California Verbal Learning Test II; SDMT: Symbol Digit Modalities Test. All reported p-values were corrected using FDR correction.

